# A naturalistic study of the efficacy and acceptability of rTMS in treating major depressive disorder in Australian youth

**DOI:** 10.1101/2024.06.20.24309219

**Authors:** Aleksandra Miljevic, Kyle Hoath, Kerry S. Leggett, Lauren A. Hennessy, Caitlan A. Boax, Jaroslaw Hryniewicki, Jennifer Rodger

## Abstract

**Objective:** Repetitive transcranial magnetic stimulation (rTMS) is an effective, evidence-based treatment for major depressive disorder (MDD) and is publicly funded in Australia. However, there is no published data to date concerning its use in private TMS service provider clinics in Australia. There is further limited data as to its efficacy and safety in treating MDD in youth populations.

**Methods:** This retrospective study examined routinely collected data of 46 outpatients aged 17 to 25 years old, who received rTMS treatment for MDD in a private TMS clinic. Primary outcomes measures were the Montgomery-Asberg Depression Rating Scale (MADRS) and the depression subscale of the 21-item Depression, Anxiety and Stress Scale (DASS-21). Secondary measures included the anxiety and stress sub-scales of the DASS-21, a measure of Quality of Life (QoL) Enjoyment and Satisfaction Questionnaire, and the Cognitive Failures Questionnaire (CFQ).

**Results:** A 4–7-week course of rTMS significantly reduce symptoms of self-reported depression (42.5% response) and clinician-assessed depression (40.7% response). Both anxiety and stress significantly reduced across the course of rTMS treatment and significant improvements to QoL and self-reported cognition were observed. Reported side effects following rTMS in youth included a mild headache and fatigue.

**Conclusions:** The findings of this naturalistic study suggest that an acute course of rTMS provided in private clinical settings is safe and effective – resulting in similar response rates in youth patients as reported in adult patients. In real world practice, rTMS proves to be a well-tolerated and highly effective intervention for MDD in youth, across diverse clinical settings.

**Implications and Contribution:** The findings of this naturalistic study suggest that in real world practice, rTMS proves to be a well-tolerated and highly effective intervention for treating depression and anxiety in youth (17–25-year-olds), with additional benefits to cognitive symptoms of depression and overall well-being.

## 1 Introduction

Depression is one of the most common mental illnesses worldwide and one of the leading contributors of global disease burden^1^. It’s estimated that 1 in 7 Australians are affected by depression and that 45% of all Australians have experienced a mental health disorder like depression within their lifetime^2^. In addition, Australia has experienced a marked increase in the prevalence of mental health issues among youth (15- to 24-year-olds)^1^. In 2007, 26% of those aged 16–24 had experienced a mental disorder within the last 12-months; in 2020–2022, this figure increased to 39%^2^, likely precipitated by the COVID-19 pandemic and its repercussions^1^.

The current pathway to seeking treatment in Australian youth includes psychotherapies usually in combination with pharmacotherapies^3^. Whilst moderately effective for most, ∼30% of adolescents and youth with depression fail to respond to pharmacotherapy treatments^4,5^, and this response is suggested to be even lower for psychotherapies, especially in moderate to severe case of MDD^6,7^. Moreover, pharmacotherapies can produce more frequent and significant side effects in youth compared to adults, including increased suicide ideation^8^. Meanwhile, Electroconvulsive Therapy (ECT) is prescribed only in rare cases due to safety concerns of seizure induction in the developing brain^9^. Overall, there are few safe, well tolerated, and effective treatment options available for youth with (severe) depression and suicidal ideation. Thus, more favourable, and effective treatment options are needed.

Repetitive Transcranial Magnetic Stimulation (rTMS) is a non-invasive, non-convulsive brain stimulation modality^10^. rTMS involves the delivery of repetitive, rapid electromagnetic pulses through the scalp to the (left or right) dorsolateral prefrontal cortex (DLPFC) via a handheld device called a ‘coil’^3^. While the exact mechanisms of action of rTMS are not yet clearly understood, high frequency stimulation to the left DLPFC and low frequency stimulation to the right DLPFC are proposed to modulate neurotransmission, synaptic plasticity and neural circuits as well as neurogenesis ^11,12^, thereby producing an antidepressant effect^13^.

rTMS is now recommended as a clinical treatment for depression by the Australian and New Zealand College of Psychiatrists^3^ and has emerged as a publicly funded, mainstream treatment in clinical practice in adults over the age of 18 years who have failed to respond to 2 or more antidepressant pharmacotherapies. For patients under the age of 18 years, no public funding is available in Australia and a secondary psychiatric opinion is required as well as the consent of both the patient and their legal guardian. In practice, however, rTMS is still predominantly applied to older adults and the results of many prominent randomised controlled clinical trials (RCTs) are based on evidence from patient populations with an average age of ∼40 years old^14–16^.

Recently, rTMS has gained traction as a treatment option in youth, with several meta-analysis and systematic reviews noting significant benefits and few side effects from rTMS application in youth patients^17–19^. In the USA, as of April 2024, the FDA has approved rTMS as an augmentative treatment option in 15+ year olds, following evidence from a large systematic review and meta-analysis from eight USA studies and 1396 patients between the ages of 8 and 24 years old^17^. At present, while rTMS is available for <25-year-olds in Australia, the effectiveness of rTMS in this population and research around safety and effectiveness is still lacking. Further, RCTs are carried out in a highly controlled environment that is not generalisable to clinical practice, where high variation in patient characteristics, medication, comorbidities, and rTMS protocols are the norm.

Research evaluating treatments in a naturalistic setting holds clinical importance as it allows for greater insight into the results in real-world clinical practice. Most Australia based naturalistic studies in large clinical services and/or hospital have focused on adult populations noting 40-58% remission rates^20–24^. Of these, only one study completed on a sample of patients from a private hospital, compared age groups including 17-15-year-olds as a comparison group (n = 33) noting higher pre-treatment depression scores and greater pre to post rTMS changes in depression compared to adults^22^.

Despite these promising results, there remains under-representation of youth specific data for both efficacy and safety information when it comes to rTMS treatment. This study aimed to further examine youth specific rTMS treatment outcomes of outpatients receiving 4-7 weeks of rTMS in a naturalistic study of a private TMS service provider clinic in Perth, Western Australia. Overall, we sought to generate novel insights from Australian population data as to the efficacy, acceptability, and safety of rTMS in the youth population. We hypothesised that rTMS treatment would significantly improve depression (both in self-reported and clinician-rated measures), anxiety, and stress in youth. We also hypothesised that rTMS would result in significantly higher self-reported quality of life and less self-reported cognitive deficits.

## 2 Methods

### 2.1 Participants and Participant Inclusion

A total of 50 patients aged 25 years or younger attended out-patient rTMS treatment at one of the nine Modalis Clinics in Perth, Western Australia between January 2021 to February 2024. Patients were included in the data set for analysis if they: (1) received rTMS for an episode of major depressive disorder (MDD; as per the Diagnostic and Statistical Manual for Mental Disorders (DSM-IV-TR; APA 2000), (2) had completed both pre-treatment and post-treatment assessment measures, and (3) had current inadequate response or intolerability to current pharmacological and/or psychological treatments. Patients with psychiatric comorbidity (i.e., anxiety disorder, personality disorder, trauma) were included, if the primary condition being treated with rTMS was an episode of MDD. Patients were excluded if they had metal in the head and/or chest, were pregnant, or presented with a current brain tumour.

All patients completed a minimum of 19 treatment sessions and both high-frequency and low-frequency rTMS outcomes were assessed at once for efficacy and safety due to the limited sample size. None of the 50 patients had received rTMS or ECT in the past and all had attempted and failed at least 2 previous pharmacotherapies for MDD. All participants, except for two, were taking antidepressants or at least one disorder related form of medication.

Given the naturalistic setting, medication changes were not exclusionary although treating psychiatrists avoided making significant concurrent medication changes during rTMS treatment, where possible. Other psychological and social therapies varied naturalistically during the rTMS treatment. A breakdown of key participant characteristics is provided in Table 1.

**Table 1.** Key patient characteristics.

|  | Youth Patient Characteristics |  |
| --- | --- | --- |
| Number of patients | 50 |  |
| Mean Age (SD) | 21.22 (±2.22) | |
| Age Range | 17 – 25 |  |
| Number and % of Females | 30 (60%) |  |
| Number and % of Males | 18 (36%) |  |
| Number and % of Transgender Individuals | 2 (4%) |  |
| Number and % patients starting on left or right rTMS treatment | Left DLPFC 10 Hz | 39 (78%) |
|  | Right DLPFC 1 Hz | 11 (22%) |

*Abbreviations:* SD = standard deviation; % = percentage of participants; rTMS = repetitive Transcranial Magnetic Stimulation; DLPFC = dorsolateral prefrontal cortex; Hz = Hertz.

The University of Western Australia (UWA) Human Research Ethics Committee provided approval for this study, and a Memorandum of Understanding was established between UWA, the Perron Institute, and Modalis for access to de-identified retrospective data.

### 2.2 Treatment protocol

rTMS was administered with a Therapeutics Goods Administration registered Neurosoft MS/D machine combined with The Neural Navigator software version 3.5 (Brain Science Tools, Utrecht, The Netherlands) for MRI-guided neuronavigation. Neurosoft figure-of-eight coils were placed on the industry standard 45-degree angle. Prior to the commencement of rTMS treatment, individual resting motor thresholds (RMT) were determined and used to prescribe rTMS treatment intensity for each individual participant by the treating psychiatrist, in accordance with standardised guidelines (RANZCP, 2018).

A total of 39 patients received left-sides high-frequency (HF) 10Hz dorsolateral prefrontal cortex (DLPFC) treatment (consisting of 75 trains of 40 pulses with 11 second intertrain intervals and a total of 3000 pulses), and 11 received right-sided low-frequency (LF) 1 Hz DLPFC treatment (consisting of 1 train off 900 pulses for a total of 900 pulses). Of these, 5 patients switched treatment, 3 from left to right DLPFC and 2 form right to left DLPFC. Left and right DLPFC stimulation sites for were localised using neuro navigational technology (guided by the patient’s own structural Magnetic Resonance Imaging [MRI] scan) for all except one patient. For this one patient, the TMS coil was placed over the left DLPFC spot which corresponds to the F3 location in the 10–20 system^25^.

rTMS stimulation intensity was adjusted in some patients who were unable to tolerate stimulation at 120% of RMT. As such, a total of 34 patients received left-sided stimulation at 120% RMT, 9 patients received right-sided stimulation at 110% RMT, 2 patients received left-sided stimulation at 110% RMT, 1 patient received left-sided stimulation at 105% RMT, 2 patients received right-sided stimulation at 90% RMT, 1 patient received left-sided stimulation at 90% RMT and 1 patient received right-sided stimulation at 60%.

Patients received an average of 34 sessions. For those patients who were deemed as having completed a course of rTMS, the minimum number of sessions was 19 and the maximum was 35 sessions. Sessions were carried out across a range of 32 to 92 days. This large range of days is mostly noted to be due to patients’ availability and unrelated (to their TMS course) illness.

### 2.3 Clinical assessments

Standardised and validated clinical assessments and outcome measures were used to assess changes to depression and secondary outcome measures. Assessments included the clinician-rated Montgomery-Asberg Depression Rating Scale (MADRS^26^ collected pre and post rTMS treatment. Further, the 21-item self-report Depression, Anxiety, and Stress Scale (DASS-21^27,28^). Both measures are standardised, robust assessments of depression, anxiety, and stress prevalence. For all, a higher score indicates greater frequency of symptoms. To assess for quality of life and life satisfaction, the self-reporting Quality of Life (QoL) Enjoyment and Satisfaction Questionnaires (QoLES-Q^30^) were used – where a higher score indicates greater life satisfaction and enjoyment. To assess for changes in cognition, the Cognitive Failures Questionnaire (CFQ^31^) was used, the CFQ is a subjective, self-report measure which asks individuals to consider the everyday mistakes they make to evaluate perception, memory, and cognition – here, a higher score indicates greater frequency of cognitive ‘failures’ (i.e., forgetfulness, failures of perception).

All measures except the MADRS were assessed pre-treatment (pre-tx.), half-way through rTMS treatment (mid-tx.), and post-treatment treatment session (post-tx.). The MADRS was assessed only at pre-tx. and post-treatment timepoints. Safety and side effects of rTMS treatment were monitored across each of the treatment sessions. The MADRS and DASS-21 depression subscale were used as the primary measure for assessing depression severity and depression response. Responders were defined as 50% or greater reduction in MADRS and/or DASS-21 depression scores from pre-treatment to post-treatment assessment.

### 2.4 Statistical Analysis

All statistical significances were set at *p*<.05 and analyses was performed using Statistical Package for the Social Sciences (SPSS) version 22.0. Means and standard deviation were calculated for continuous variables, and frequency and percentage for all other variables. Linear Mixed Effects Models (LMM) fitted with restricted maximum likelihood estimations were used to determine changes in depression (MADRS, and DASS-21 depression subscale), anxiety and stress symptoms (DASS-21 subscales), as well as across QoL (QoLES-Q) and cognition (CFQ) across the pre-rTMS, mid-rTMS, to post-rTMS treatment period. This statistical method enables the assessment of changes over time (i.e., repeated measures) and is robust to violations of normality ^32^. LMMs provide accurate, unbiased estimates irrespective of missing data, and are capable of flexibly capturing skewed and non-normal distributions to identify robust models of data despite limited sample size. The F-test was used to evaluate if there were between-group differences and test for interactions across time.

Where significant differences were revealed, additional post-hoc comparisons using a paired-samples t-tests to assess pre-tx. to mid-tx., pre-tx. to post-tx., and mid-tx. to post-tx. differences were conducted to reveal the specific changes across the three time points.

Additional Pearson’s *r* correlations were conducted to investigate the relationship between baseline depression in relation to anxiety, stress, QoL, and cognition, as well as the relationship between post-treatment depression in relation to anxiety, stress, QoL, and cognition.

## 3 Results

Of the 50 youth patients that undertook treatment at Modalis, 4 individuals were excluded from the analysis as there was not enough data to include them in analysis (i.e., data from a minimum of 2 timepoints was missing, due to missed appointments or patients not completing the outcome measures). Thus, a total of 46 individuals were included in the final analysis.

### 3.1 Safety, tolerability, and reported side effects of rTMS

A total of 24 patients (52.17%) reported experiencing mild fatigue as a result of rTMS treatment and 15 continued to experience fatigue in the latter half of treatment but not severely enough to cease treatment. Further, 20 patients (43.48%) reported mild headaches due to rTMS treatment within the first 1-15 sessions. Of these, 6 patients continued to experience headaches in the latter half of treatment but not at every session and not severely enough to cease treatment.

Other reported side effects included light-headedness (4.45% of patients) and mild nausea (2.17% of patients). There was a single case of suicide ideation however, this was not attributed to the rTMS treatment as the individual was reporting suicidal thoughts prior to the commencement of treatment. Overall, rTMS was well tolerated despite instances of participants having experienced mild fatigue and headaches.

### 3.2 rTMS-related changes in depression

The LMM revealed significant reductions in depressive symptoms over time as assessed on the self-report DASS-21 sub-scale, *F*(1, 44) = 227.14, *p*<0.001, as well as clinician-rated reductions in the MADRS, *F*(1, 71) = 534.306 *p*<0.001.

A total of 40 self-report DASS-21 depression sub-scale were available and of these, a total of 17 (42.5%) individuals responded to rTMS based on 50% or greater reduction in scores. Meanwhile, 15 patients (37.5%) reported a decrease in symptoms ranging from 8 – 39% improvement and 8 patients (20%) did not benefit from rTMS at all. Further, a total of 27 MADRS rating scale scores were available with 11 patients (40.74%) demonstrating 50% or greater reduction in scores, 12 patients (44.44%) noting 8 – 44% improvements, and 2 patients (7.41%) demonstrating no improvement to depression scores. Overall, this indicates that rTMS treatment significantly decreased and thus improved depression in youth patients (Figure 1).

**Figure 1.**
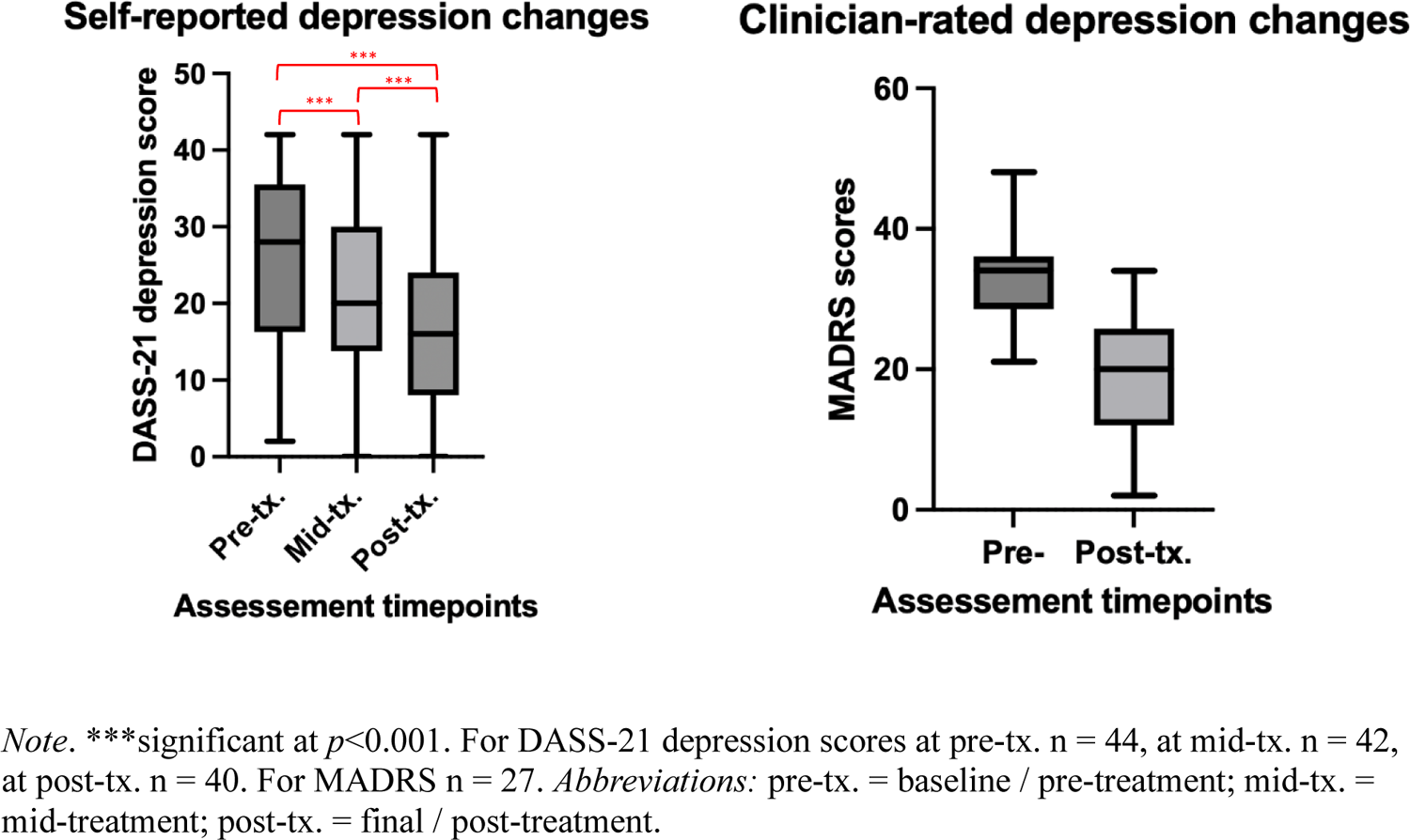
Self-report and clinician rated changes in depression before, during and after rTMS treatment in a youth population.

### 3.3 rTMS-related changes in anxiety and stress

The LMM revealed significant reductions in self-reported anxiety symptoms over time as assessed on the DASS-21 sub-scale, *F*(1, 45) = 130.70, *p*<0.001. Further, a significant reduction in self-reported stress as assessed on the DASS-21 sub-scale was also revealed, *F*(1, 45) = 276.98 *p*<0.001. Overall, this indicates that rTMS treatment significantly decreased and thus improved anxiety and stress in youth patients (Figure 2).

**Figure 2.**
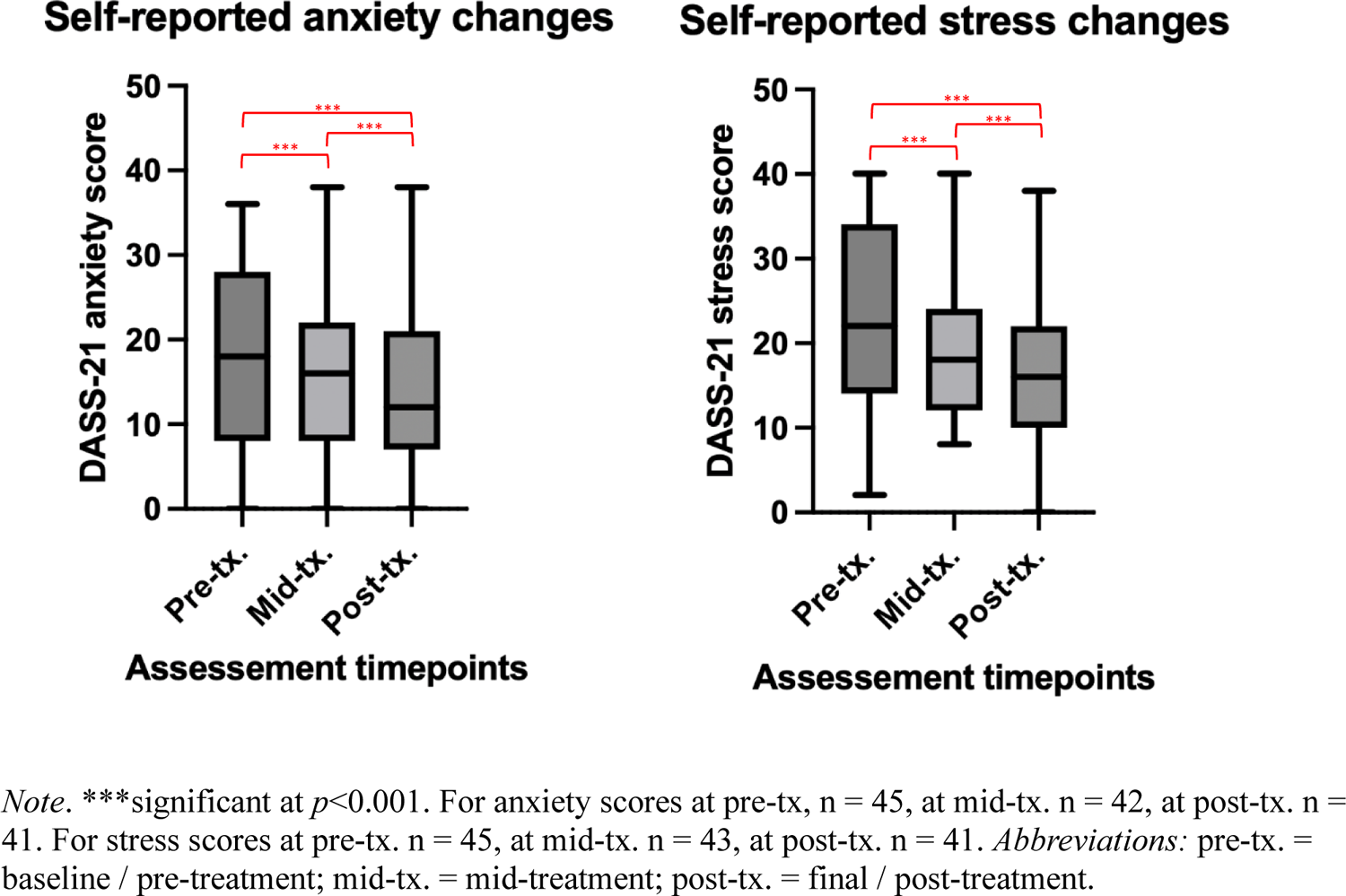
Self-report and clinician rated changes in anxiety and stress before, during and after rTMS treatment in a youth population.

### 3.4 rTMS-related changes in quality of life and cognition

The LMM revealed significant increases in self-reported QoL symptoms over time as assessed on the QoLES-Q, *F*(1, 46) = 702.88, *p*<0.001. Overall, this indicates that rTMS treatment was able to significantly increase QoL in youth patients. Additionally, the LMM revealed significant decreases in self-reported cognitive ‘failure’ symptoms over time, *F*(1, 45) = 449.13, *p*<0.001. Overall, this indicates that rTMS treatment significantly improved self-reported cognitive mistakes in youth patients (Figure 3).

**Figure 3.**
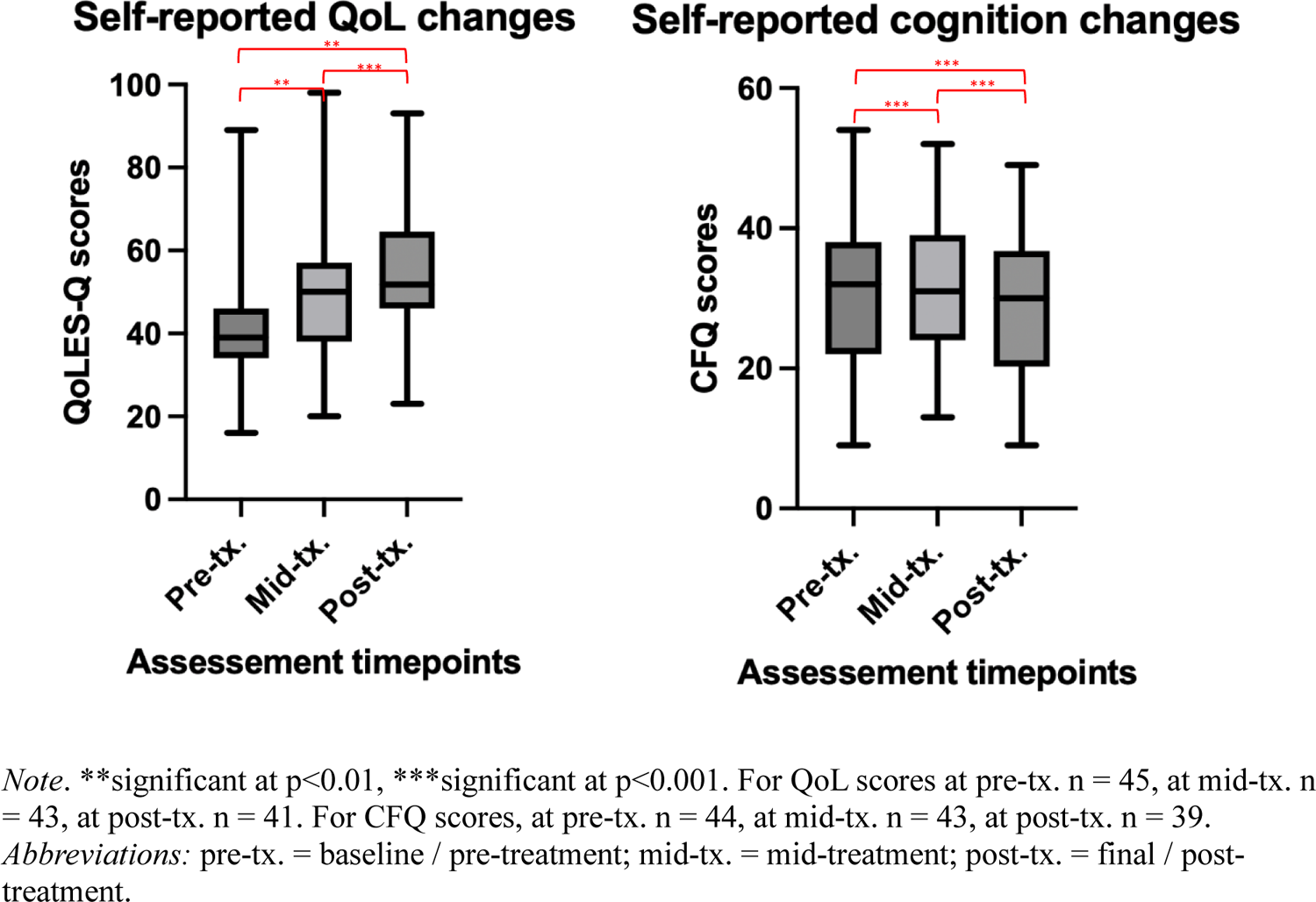
Self-report and clinician rated changes in QoL and cognition before, during and after rTMS treatment in a youth population.

### 3.5 Relationships between depression, and anxiety, stress, QoL, and cognition

Significant positive relationships were observed between baseline depression (based on the DASS-21 but not MADRS) with both anxiety (*r* = .56, *p*<0.001) and stress (*r* = .71, *p*<0.001). Further, baseline depression (on DASS-21 but not MADRS) was significantly and negative correlated to lower baseline QoL (*r* = −.32, *p*=0.03) but not with cognitive failure symptoms (Figure 4).

**Figure 4.**
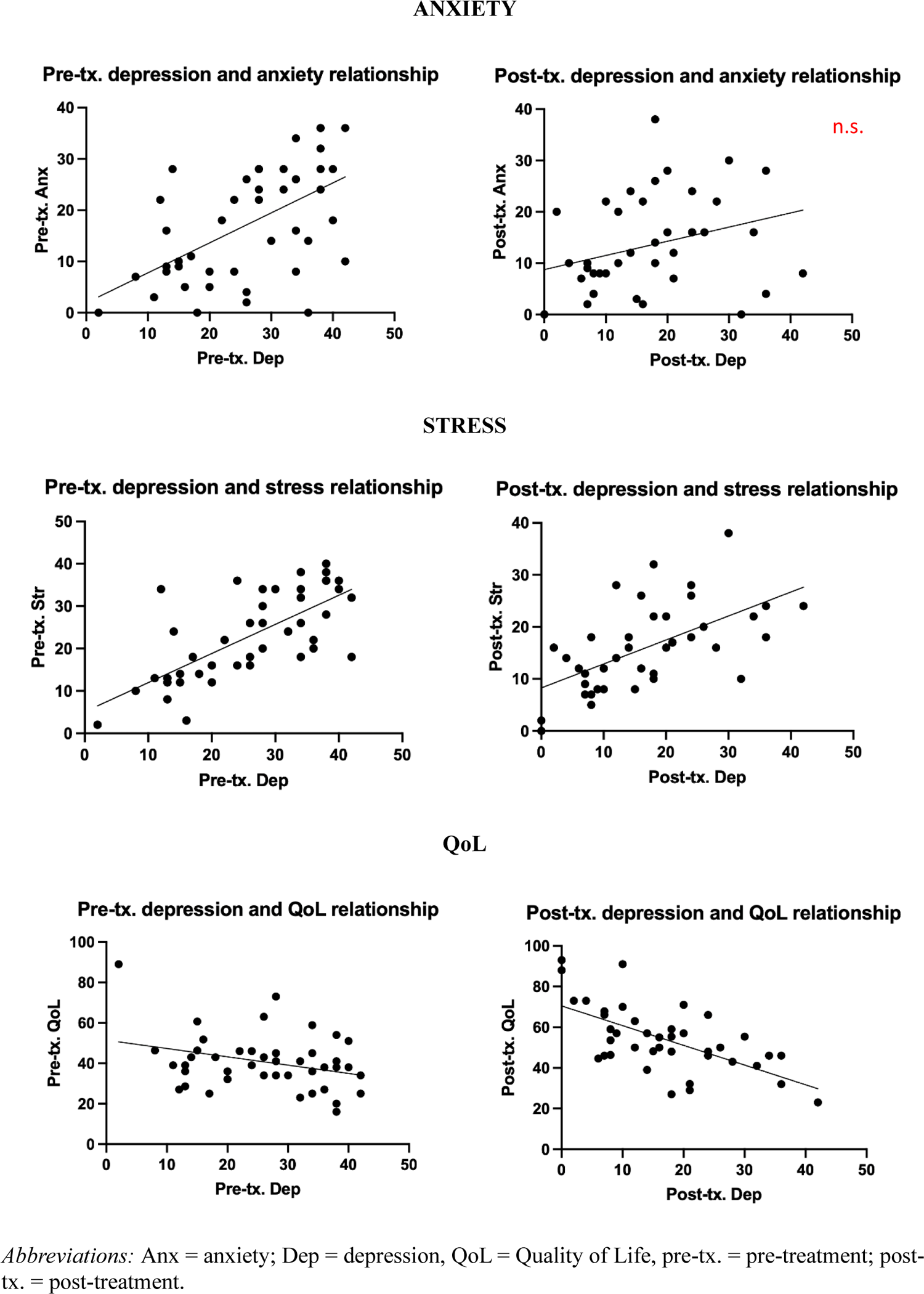
Correlational assessment of the relationship between pre and post treatment depression scores with anxiety, stress, QoL, and subjective cognition.

In addition, post-treatment depression based on the DASS-21 was significantly positively correlated with stress (*r* = .59, *p*<0.001) but not anxiety. Further, post-treatment depression based on DASS-21 was significantly, negatively correlated with post-treatment QoL (*r* = .63, *p*<0.001) but not with cognitive failure symptoms (Figure 4). Interestingly, post-treatment MADRS was also significantly negatively correlated with self-reported post-treatment anxiety (*r =* .55, *p*=0.004), stress (*r =* .55, *p*=0.004), QoL (*r* = .55, *p*=0.004) and cognitive failure symptoms (*r* = .40, *p*<0.05).

## 4 Discussion

To the best of our knowledge, this is the first naturalistic study in Australia to assess the acute rTMS treatment outcomes from an out-patient private TMS service provider clinic, in youth presenting with a current MDD episode. As hypothesised, our study found a significant improvement in self-reported and clinician-rated depression in youth who received rTMS treatment. Overall, 80% of patients self-reported a benefit from rTMS treatment. Specifically, the response rate (defined by a 50% reduction in depression scores) of 42.5% appears to be mostly consistent with the range of response reported in other studies assessing rTMS efficacy in both adults (∼46–68%)^33,34^ and youth (∼41–56%)^35,36^. However, our observed response rates may be considered slightly lower than in previous published works on youth, likely due to the naturalistic nature of our study and the less controlled, more varied treatment parameters and clinical characteristics of the included patients.

The study revealed that rTMS treatment was generally well tolerated with some transient mild headaches and fatigue, as well as rare occurrences of light-headedness. This is encouraging, and consistent with side-effects reported in other TMS research in adults^37,38^ and in youth^17–19^. There were no documented seizures or increases in suicide ideation due to rTMS treatment. Most side effects appeared to significantly improve or resolve after the first two weeks of treatment and were not influential enough to warrant cessation of treatment by the patient or treating psychiatrist.

It has been estimated that approximately 65-85% of patients with depression also experience symptoms of anxiety^39,40^. In addition to significant reductions in depression, this study revealed that comorbid anxiety and stress also improved post-treatment. This is consistent with the literature in pharmacological^41^ and rTMS studies^42–44^, and in naturalistic^20–24^ and clinical trial^45,46^ studies, where improvements in depression are often accompanied by improvements in anxiety. Within this youth sample, a reduction in anxiety was observed in 69% of participants, somewhat higher than what is reported in the literature (∼48%-61% of participants improved^43,45^).

Further, we found that pre-treatment depression significantly related to pre-treatment anxiety, but this relationship was no longer significant at the post-treatment timepoint. If anxiety and depression are highly correlated, as the literature suggests, we would expect their relationship to be maintained even after treatment. While this was the case for MADRS and the DASS anxiety sub-scale, there was no significant relationship between self-reported depression and anxiety on the DASS post-treatment. One reason for a lack of significant relationship at this post-treatment timepoint may be that rTMS is having more effect on one disorder than the other. However, the therapeutic mechanisms of rTMS in depression *and* anxiety remain poorly understood^47,48^. Alternatively, the different results in clinician-rated and self-report post-treatment measures could be due to variation in perception of symptoms between patient and clinician^49^. The increase in the variability and spread of reported symptoms at the post-treatment timepoint for both depression and anxiety may be due to the small sample size or may reflect individual differences in symptoms and symptom presentation. Nonetheless, there is a clear effect of rTMS on both anxiety and depression in youth within this sample.

Interestingly, self-reported stress improved as a result of rTMS, consistent with evidence for positive effects of rTMS on disorders such as post-traumatic stress^50^. However, the specific dimension of stress assessed within the DASS scale is perceived stress, which is characterised primarily by symptoms of tension, inability to relax, irritability, fidgety or jumpy, and/or easily upset, symptoms which overlap and are often associated with anxiety, as well as situational / external demands. Thus, while rTMS may be treating stress, it is also possible that the drop in stress could be attributed to reduced stress around seeking treatment / receiving the rTMS intervention rather than the rTMS intervention itself. There was also a significant, positive relationship between stress and depression that was maintained at both the pre- and post-treatment time points, as has been reported for antidepressant medication treatment^51^. Depression, anxiety and stress are highly correlated^52^ suggesting a need for future research to investigate these disorders independently, and their responsiveness to rTMS treatment.

Our findings further indicate that the reduction of depressive symptoms following rTMS is accompanied by an improvement in QoL. Specifically, our results show a significant increase in QoL as well as a persistent negative correlation between both pre- and post-rTMS depression and pre- and post-rTMS QoL, respectively. Thus, lower levels of depression related significantly to higher levels of QoL. This finding is supported by broad evidence for a relationship between improvements in depression (no matter the treatment type) and QoL^55–58^. This is an important observation, indicating that effective treatment can induce an improvement in not only depression but also overall well-being in a relatively short timeframe (i.e., 4-7 weeks).

Approximately 40% of patients with depression have been found to experience cognitive impairments^53^. In our sample, rTMS improved subjective cognition, which aligns with the results of meta-analyses noting modest effect size improvements in cognition in active as compared to sham rTMS treatment^59,60^. While some studies assessing different treatment modalities report a correlation between improvements in self-reported cognition and depression symptoms^54^, this is not a consistent effect^53^, and not one we see in our youth sample. However, many of the published results pertaining to cognition, depression, and the effects of different treatments, are based on standardised tests of cognition and cognitive control, whereas we assessed subjective, self-report measures. In addition, it may be that a larger sample size is needed to detect robust, significant differences in cognition. Future research should aim to further explore the relationship between self-report measures of cognition and symptoms of depression.

Limiting our analysis, certain details around patient history (e.g., duration of episode, diagnosis date etc.) were not available as this data was extracted from an outpatient rTMS service provider and de-identified records. In addition, due to missing data, some of the comparisons (i.e., MADRS pre- to post-treatment) may be somewhat underpowered. Not all patients completed the full recommended 35 sessions, and not all completed treatment within the recommended 4-7 weeks. For some, treatment took as long as 13 weeks, due to patient-related life circumstances. Further, while patients were allocated across three rTMS protocols (HF-left DLPFC, LF-right DLPFC, or sequential bilateral rTMS), due to the naturalistic design and limited sample size, we were unable to perform comparisons between the different protocols. Notwithstanding these limitations, the strength of this study is the significant clinical outcome data collected in a naturalistic setting, which reflects real world practice, where high rates of psychiatric comorbidity, varied current medication use, and treatment resistance is the norm.

Future research should aim to assess and report on larger youth patient populations, as well as compare efficacy of different rTMS treatment protocols. Additionally, it will be important to compare youth response rates to adult response rates in naturalistic clinical settings. Finally, our results highlight the need to standardise and incorporate measures of QoL and cognition into rTMS research and clinical practice to better assess patient outcomes.

## 5 Conclusion

The findings of this naturalistic study suggest that an acute course of rTMS provided in a private clinical setting results in similar response rates to the existing rTMS literature in the youth population. Further, our study notes significant improvements in anxiety, stress, QoL and cognition, indicating that rTMS treatment can produce an improvement in overall well-being in a relatively short timeframe. This study adds to the growing body of research showing that rTMS is an important therapeutic option in real-world practice for treating MDD in Australian youth. Further clinical implementation and research into the efficacy of rTMS in youth populations is warranted.

## Data Availability

All data produced in the present study may be available upon reasonable request to the authors.

